# Framing Major Depressive Disorder as a Condition of Network Imbalance at the Compartment Level: A Proof-of-Concept Study

**DOI:** 10.1101/2024.09.23.24314178

**Authors:** Tien-Wen Lee

## Abstract

**Background:** Hypoactivity in the frontoparietal (FP) system and hyperactivity in the limbic system have been observed in major depressive disorder (MDD). Among the neuropathological theories of MDD, the cortico-limbic dysregulation model has been prominent, and it was recently extended and refined by the framework of imbalanced reciprocal suppression between the FP and limbic compartments. This research aims to examine this refined theory.

**Methods:** Sixty MDD and sixty healthy control datasets, including structural magnetic resonance imaging and resting-state functional MRI (rsfMRI), were selected from the CAN-BIND database. The cerebral cortex in rsfMRI was parcellated by MOSI (modular analysis and similarity measurements). For each parcellated node, the mean amplitude of low-frequency fluctuation (mALFF) was computed. Correlation analyses were employed to construct an adjacency matrix between partitioned nodes and to examine the relationship between nodal power and nodal degree.

**Results:** Two-sample t-tests revealed that the limbic system in MDD was associated with a higher partition number and mALFF (p < 0.005). A negative correlation between nodal degree and nodal power was replicated (p < 1E-10), indicating that higher functional input may more effectively suppress regional neural activity. For FP nodal power and Limbic-FP connectivity, the MDD group exhibited a more negative correlation (stronger suppression), while for limbic nodal power and FP-Limbic connectivity, the MDD group showed a less negative correlation (weaker suppression).

**Conclusions:** Consistent with the inter-compartmental network imbalance framework, MDD is characterized by abnormal cortical signal organization and aberrant reciprocal suppression between the FP and limbic systems.

## Introduction

Major Depressive Disorder (MDD) is a prevalent and disabling condition that imposes a significant burden on patients, their families, and society (Ferrari et al., 2013; Vos et al., 2015). In the United States, the 12-month prevalence of MDD is estimated to be 7%, with females experiencing rates 1.5 to 3 times higher than males, beginning in early adolescence (American Psychiatric Association, 2013). While the core symptoms include depressed mood and loss of interest, along with social and occupational impairment, MDD manifests a broad array of additional symptoms. These include cognitive decline (“pseudodementia”), fatigue, feelings of guilt or worthlessness, psychomotor agitation or retardation, sleep disturbances (insomnia or hypersomnia), appetite changes, significant weight fluctuation, suicidal ideation, and even psychotic symptoms. Furthermore, MDD is often comorbid with other psychiatric disorders, such as anxiety disorders, obsessive-compulsive disorder, post-traumatic stress disorder, and substance dependence (Oquendo et al., 2005; Kessler et al., 2008; Brière et al., 2014), suggesting an underlying pathology that increases susceptibility to various conditions.

Consistent with its clinical presentation, brain abnormalities in MDD are widespread, affecting both cortical regions and subcortical nuclei (Drevets, 1999; Wu et al., 1999; Mayberg et al., 2000; Kennedy et al., 2001; Sheline et al., 2001; Drevets et al., 2002; Mayberg, 2003; Holthoff et al., 2004; Mayberg et al., 2005; Aihara et al., 2007; Siegle et al., 2007; Hwang et al., 2010; Smith et al., 2011a; Elliott et al., 2012; Ruhe et al., 2012; Lee et al., 2013; Phillips et al., 2015; Pilmeyer et al., 2022). This distributed pattern of neural dysfunction suggests that MDD is unlikely to be caused by a single abnormal brain region or circuitry, but rather by the malfunctioning of large-scale networks. In line with this, a consistent neural pattern in MDD has been identified—hypoactivity in the frontal (or frontoparietal; FP) network and hyperactivity in the limbic (LM) system (Drevets, 1999; Mayberg, 2003; Pilmeyer et al., 2022). This observation supports the limbic-cortical dysregulation model proposed by Mayberg (Mayberg, 1997), which has since been refined and expanded, forming the central focus of this study (Lee, 2016; Lee and Xue, 2018b).

### Rationale for Conceptualizing MDD as an Imbalance Between Limbic and Frontoparietal Compartments

There is strong evidence of limbic hyperactivity and hypofrontality (fronto-parietal hypoactivity) in MDD. Limbic hyperactivity has been observed in the anterior cingulate cortex (ACC), posterior cingulate cortex (PCC), OFC, amygdala, and parahippocampal gyrus during the acute phase of MDD (Kennedy et al., 2001; Drevets et al., 2002; Aihara et al., 2007; Drevets et al., 2008). In addition, amygdalar disinhibition and exaggerated activation were exhibited in response to cognitive demand and subliminal facial emotions (both positive and negative) (Sheline et al., 2001; Lee et al., 2013). On the contrary, clinical improvement is accompanied by metabolic and response normalization in the limbic system (Wu et al., 1999; Mayberg et al., 2000; Kennedy et al., 2001; Sheline et al., 2001; Drevets et al., 2002; Holthoff et al., 2004). Note that this proposal uses the term “limbic system” to represent a broader concept that hosts the midline, outer and deep cortical portions of the default-mode network (DMN) (Lee and Xue, 2018a).

Hypofrontality, another pathognomonic feature of MDD, was noticed in the dorsolateral prefrontal cortex (dlPFC) and the medial PFC (mPFC) (Kennedy et al., 2001; Drevets et al., 2002; Mayberg et al., 2005; Aihara et al., 2007; Siegle et al., 2007). In addition, during the depressive state in MDD, reduced frontopolar activities to both positive and negative social stimuli and to cognitive challenges were observed (Elliott et al., 2012). Remission is associated with normalizing hypo-metabolism in several frontal regions (Mayberg et al., 2000; Kennedy et al., 2001; Drevets et al., 2002). The re-emergence of depressive symptoms in remitted MDD that followed short-term discontinuation of specific serotonin reuptake inhibitors (SSRIs) was accompanied by a decrease in regional cerebral blood volume in the left mPFC, which was correlated with the symptom severity (Henry et al., 2003). In either remitted or acute depression, the induction of sadness was tied to decreased blood flow in the mPFC (Liotti et al., 2002). Transcranial magnetic stimulation over dlPFC and mPFC may enhance frontal function and improve depression (Speer et al., 2000; Downar et al., 2014). Although the superior and inferior parietal cortices have attracted less attention in MDD research, their decreased activities are often in tandem with those in the PFC (Disner et al., 2011; Pimontel et al., 2016). Cerebrovascular lesions in the white matter may unfavorably impact the operation in the FP compartment and lead to secondary depression (Culang-Reinlieb et al., 2010; Santos et al., 2010). A recent review verified the above-summarized patterns consistently observed for two decades (Pilmeyer et al., 2022).

Notably, the findings from independent brain research of MDD are distributed broadly in the PFC, parietal cortex, and limbic region. The involved circuitries comprise but are not limited to emotion regulation, emotion processing, reward, saliency, executive control (Brand et al., 2015; Phillips et al., 2015; Pimontel et al., 2016), etc. The abnormality of single or few circuitries cannot accommodate the multi-faceted symptomatology of MDD (Chen et al., 2005; Heinzel et al., 2009; Hwang et al., 2010; Lee et al., 2011; Busatto, 2013; Bagherzadeh-Azbari et al., 2019). They altogether decline the conjecture that the pathological foci of MDD are localized/confined or attributable to limited circuitries but indicate an imbalance between the limbic system and the FP portion of the cortical mantle. The insight is paramount since it suggests a compartment-level dysfunction to account for MDD, which signifies a possible fundamental difference between psychiatric and neurological conditions: most neurological disorders involve discrete and localizable pathology, such as vascular events, tumors, degenerative changes (in the early phase), epileptogenic foci, demyelinating alterations, etc. MDD pathology, as a canonical example of psychiatric disorders, may engage two (or more) large networks (Lee, 2016; Lee and Xue, 2018b). “Compartment” is henceforth used to represent the neural substrates that embraced much broader brain regions (large network), in contrast to circumscribed circuitry/network (e.g., saliency circuitry, DMN) used in extant literature. Even though the insightful cortico-limbic dysregulation theory was proposed two decades ago, the exploration that takes the whole FP and limbic systems as processing units is still pending.

### Analytic framework to examine compartment-level pathology

Although the issue looks straightforward, an innate difficulty must be overcome to enable the assessment. Current fMRI may record neural information at a spatial resolution of several millimeters, allowing the brain to be divided into tens to hundreds of thousands of voxels. While enhanced resolution is a favorable development, the abundance of voxels presents an obstacle in analyzing brain networks. As the temporal dynamics between neighboring fMRI voxels are not independent, investigators have applied dimension reduction strategies to simplify the data structure. One common approach for cortical partition is to divide the brain into distinct regions based on anatomy, although its validity has frequently been questioned (Amunts et al., 1999; Smith et al., 2011b; Wig et al., 2011; Stanley et al., 2013; Lee et al., 2014; Xue et al., 2014). It would be ideal to partition the brain dynamics based on fMRI signal itself, i.e., functional parcellation (FCP), in contrast to relying on the atlas-informed structural parcellation (Shi and Malik, 2000; Damoiseaux et al., 2006; Smith et al., 2009; Shen et al., 2013). I developed an innovative analytic scheme—Modular Analysis and Similarity Measurements (MOSI)—to address this issue (briefly introduced in Methods) (Lee and Tramontano, 2021). MOSI is a data-driven algorithm that requires no additional premises and enables individualized, multi-resolution parcellations. By segmenting resting-state fMRI (rsfMRI) data into functionally homogeneous clusters, MOSI laid the foundation for the analytics in this research. Each cluster, composed of voxels with similar BOLD signal traces, is designated as a neural node, and the communication between these nodes defines a network.

Following MOSI, the mean power, represented by the amplitude of low-frequency fluctuations (ALFF), was calculated for each node. The interaction between nodes was quantified using the degree of correlation between their mean time series, also known as functional connectivity (FC). In addition, the negative relationship between regional and inter-regional profiles was examined based on my recent work (Lee and Xue, 2017b; Lee and Tramontano, 2024). It is expected that the interaction between FP and limbic compartments is deviated in MDD. Comparisons between MDD and normal groups can be categorized into three domains: within the FP compartment, within the limbic compartment, and notably, between the FP and limbic compartments, see Figure 1.

**Figure 1.**
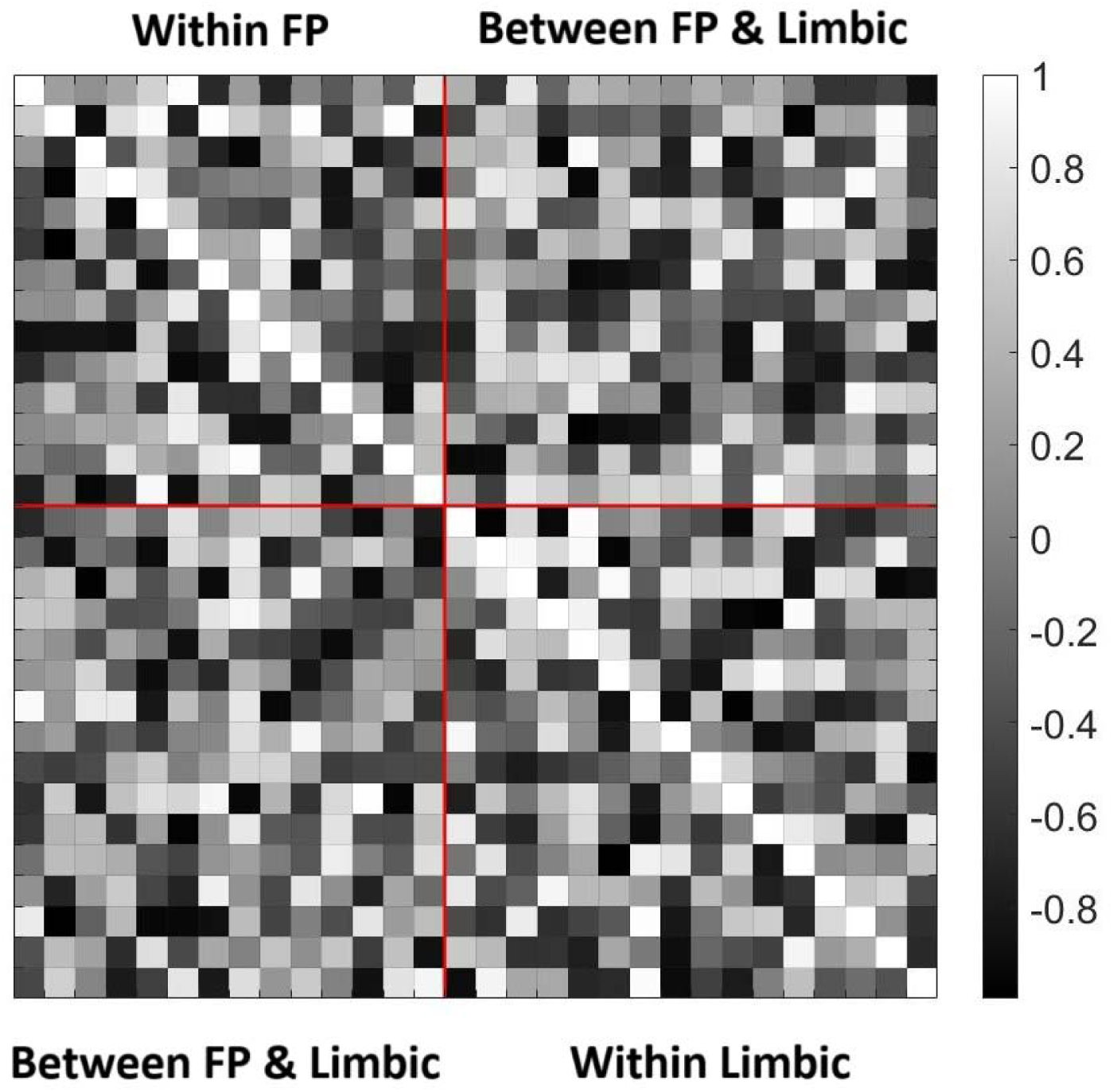
An illustration of the three categories of analyses using a tentative correlation matrix: within the FP compartment (left upper), within the limbic compartment (right lower), and between the FP and limbic compartments.

The Desikan-Killiany atlas delineates anatomical regions belonging to the FP and limbic systems (Desikan et al., 2006). For the limbic system, Mega et al.’s definition, distinguishing the OFC/amygdala-centered paleocortical division and the hippocampus/cingulate-centered archicortical division, were followed (Mega et al., 1997). Each MOSI’s partitions can then be categorized into FP, limbic, or other brain lobes. Based on the understanding that the DMN carries limbic information flow (Lee and Xue, 2018a), the middle temporal and inferior parietal cortices were also categorized as part of the limbic system in terms of functional (not anatomical) attributes. By fulfilling functional parcellation, MOSI offers flexibility regarding the spatial extent of functional units (modules) while adhering to brain science knowledge.

## Material and methods

### Subjects, MRI data, and preprocessing

Baseline MRI data were collected from a total of 120 subjects, including 60 patients with MDD and 60 healthy controls, all sourced from the Canadian Biomarker Integration Network in Depression (CAN-BIND) (Lam et al., 2016), in which written consent was obtained from each participant. Subjects with incomplete demographic information, insufficient clinical evaluations, or suboptimal imaging quality (i.e., too much movement, poor segmentation) were excluded. Private Pearl IRB (www.pearlirb.com) approved this study and issued a certificate of ’Exempt Research Determination’ (IRB ID: 2023-0133) at the request of CAN-BIND. Functional and structural MRI (sMRI) images of the whole brain were acquired using 3.0 Tesla MRI scanners (GE, TR=2s, voxel size=4×4×4 mm³, 300 volumes). For detailed information about the scanning procedures and study protocol, please refer to (Lam et al., 2016).

The echo planar imaging (EPI) data were processed using the Analysis of Functional NeuroImages (AFNI) software package (Cox, 1996). Preprocessing steps for rsfMRI included despiking, slice-time correction, realignment (motion correction), registration of T1 anatomy, spatial smoothing, and bandpass filtering at 0.01–0.15 Hz (Tong et al., 2019). The first 4 scans were discarded. A smoothness kernel of 8 mm was applied to the EPI data. The FreeSurfer software was used to segment gray and white matter from T1-weighted sMRI data and to parcellate the cortical mantle into 68 regions of interest (ROIs) based on the Desikan-Killiany Atlas (Dale et al., 1999; Fischl et al., 1999; Desikan et al., 2006), providing the initial partition for subsequent MOSI analysis. Following the analytic pipeline developed by Jo et al. (Jo et al., 2010; Lee et al., 2014; Xue et al., 2014), 12 movement parameters, along with white matter and ventricular signals, were modeled in the regression (Jo et al., 2010). Additionally, third-order polynomials were used to fit baseline drift. ALFF was calculated as the square root of the band-passed power of BOLD dynamics. Pearson correlation coefficient (CC) was computed pairwise between the mean temporal traces of any paired voxels or clusters to indicate FC.

### MOSI analysis and adjacency matrix construction

The connectivity map based on CCs is the foundation for modular analysis. Modules or communities are groups of nodes within a network that are more densely connected to one another than to other nodes, as defined by the CCs. MOSI incorporates modular analysis and similarity measurements, splitting a module into sub-modules and uniting similar sub-modules into a bigger module. MOSI fulfills the parcellation of the cortex abiding by only two criteria: neighborhood and similarity, with adjacent voxels sharing similar neural activities grouped together, precisely the rationale behind FP of cortical signals. The splitting-unifying processes were repeated until convergence. The current version of MOSI uses the Louvain community detection algorithm. The number of modules and, hence, the partition resolution can be titrated by gamma values, with a higher value yielding more modules (Lee and Tramontano, 2021). We explored gamma values ranging from 0.45 to 0.90 (10 different resolutions). The unique features of MOSI include: (1) it is an individualized analysis feasible for the heterogeneity in MDD; (2) it lays out a plausible foundation for subsequent network analysis; (3) it enables a multi-resolution approach to investigate brain informatics at different scales (Lee and Tramontano, 2021).

With MOSI-defined partitions and corresponding neural nodes, an adjacency matrix can be constructed by first calculating the correlations between the mean time series of each node pair, and these CCs are then converted to z-scores using Fisher’s transformation (denoted as zCCs). Given that the module sizes derived by MOSI vary, it is crucial to apply appropriate weighting strategies to the resulting adjacency matrix. Assume there are N nodes, and nodes i and j have size Ni and Nj, respectively. In terms of connectivity, the connection strength between nodes i and j can be weighted 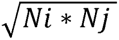, then normalized by average weight across all possible connections excluding the diagonal components (weighting strategy I).

### Statistical comparisons

#### MOSI results analysis

For each gamma value and the associated partition, two-tailed, independent two-sample t-tests were performed between MDD and Normal groups regarding the partition number, mean voxel number per partition, and mean ALFF, with a null hypothesis that there were no between group differences. Since spatial extents of the MOSI results were not fixed and varied with different gamma values, the total voxel number and ALFF were also compared. The statistical thresholds were set at p-values < 0.01 and < 0.005 to reflect different levels of stringency. QQ-plots were employed to explore differences in module size distributions between the MDD and Normal groups. If differences are confirmed, post-hoc analyses, as described above, will be conducted to clarify the effects of size on neural metrics. Several indices that are not central to the main focus of this study are summarized in the Supplementary Materials (Part IV), including the mean CCs of the FP system, limbic system, between the FP and limbic systems, and overall mean CCs (combining the FP and limbic compartments) and their associated integrative graph indices (Rubinov and Sporns, 2010).

#### Exploring the relationship between power and connectivity

The investigation of regional and inter-regional relationships is driven by the observation that global connectivity of a node (i.e., nodal degree in graph theory, defined as the summed connection strengths centered on a node) may influence the power of that specific node (Lee and Xue, 2017b; Lee and Tramontano, 2024). It has been noted that nodal degree at the lower end of the spectrum of neural signals (fMRI and delta/theta spectrum in EEG) may reflect more inhibitory information, with a higher nodal degree robustly correlating with reduced nodal power (ALFF) (Lee and Tramontano, 2024). Importantly, while correlation analysis is typically bidirectional, in this context, the relationship assumes certain degree of directionality: It is the nodal degree that may influence nodal power (from all-to-one perspective), less likely vice versa (one-to-all counterpart). This assumption is supported by two arguments: (1) Similar to FC, a negative relationship between nodal degree derived from structural connectivity (SC) and ALFF has also been observed (Lee and Xue, 2017b). Since the underlying hardwiring dictates the emergent property—indicating that SC influences local power not the reverse—the fact of a substantial correspondence between FC and SC suggests that the causal inference can similarly be extended to the FC scenario (Skudlarski et al., 2008; Honey et al., 2009; Lee and Xue, 2018c). (2) Consider a system that has nodes ranked by ALFF magnitude, with power driving connectivity in a negative direction. Given the negative relationship, the node with the lowest ALFF is expected to have the weakest connections (nodal degree) from the remaining nodes with higher ALFFs. However, this creates a contradiction, as the negative relationship suggests that a lower nodal degree should result in higher ALFF. This reinforces that nodal degree influences nodal power, rather than the opposite. It is expected that the slope relating the regional power and inter-regional functional connectivity is altered in MDD. Referring to Figure1, three classes of power-connectivity relationship are to be explored. (1) FP-Limbic: the FP nodal ALFF influenced by the connections between FP and limbic systems; (2) Limbic-FP: the limbic nodal ALFF influenced by the connections between FP and limbic systems; and (3) the overall relationship between nodal power and connection combining FP and limbic systems. Class (3) is used to examine whether the negative relationship between nodal power and nodal degree can be replicated in these data samples, while classes (1) and (2) are the primary focus for uncovering insights into FP and limbic interactions.

Operationally, the nodal power (mean ALFF over all voxels within a particular partition) and mean nodal degree for all nodes of each individual were concatenated to enable group-level analysis of the power-connectivity relationship. Before concatenation, the individual data was standardized by subtracting the mean and dividing by the standard deviation to facilitate valid comparisons. The statistical examination of the difference between power-connectivity CCs for MDD and Normal groups was computed as follows: Assume the CCs for MDD and Normal groups are r1 and r2, with data number N1 and N2. The statistical z-score, denoted as zDiff, can be calculated using the formula (atanh(r1)-atanh(r2))/sqrt(1/(N1-3)+1/(N2-3)), where atanh and sqrt indicates hyperbolic arctangent function and square root function, respectively. The p value is then computed as 2*(1-normcdf(abs(zDiff))), where normcdf is a MATLAB function (R2023a; The MathWorks, Inc., Natick, Massachusetts, United States) that calculates the cumulative distribution function of the normal distribution, providing the probability that a normally distributed random variable is less than or equal to a specified value.

Special consideration is required for the weighting strategy in this power-connectivity scenario. The homogeneity in brain signals within a module could be mediated and maintained by intra-modular interactions (Freeman et al., 1997; Chialvo, 2010), so it appears more reasonable to weight the connectivity based solely on the “inputs” from node j to node i by 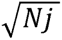, where j ∼= I, then again normalized by average weight (weighting strategy II), to explore the relationship between the nodal power and average nodal degree. Both weighting strategies were adopted, with the results based on weighting strategy II reported in the main text and the other in the **Supplementary Materials** (part II).

## Results

The MDD group had a mean age of 34.1 years (SD = 12.9), while the Normal group had a mean age of 32.7 years (SD = 11.6), with no significant difference (p = 0.552). The mean Montgomery-Åsberg Depression Rating Scale (MADRS) score for the MDD group was 29.7 (SD = 5.2), which was significantly higher than the Normal group’s mean score of 0.7 (SD = 1.6), with an extremely significant p-value (< 1E-10). Gender distribution did not differ significantly between the two groups, with 22 males and 38 females in the MDD group compared to 21 males and 39 females in the Normal group (χ² = 0.302, p = 0.583).

### MOSI outputs and the ensuing neural metrics analyses

Concordant with the nature of MOSI, the cluster number increased, and the number of voxels in each cluster decreased with the gamma values. For Normal group, the segmented module numbers of the cortex were 85.8 and 152.6 at gamma 0.45 and 0.90, respectively, with the module z-scores increasing from 0.54 (CC 0.50) to 0.75 (CC 0.64), see top subplot in Figure 2. The module number of the initial partition based on the Desikan-Killiany Atlas was 68, with an average voxel number of 129.5 (64mm^3/voxel) and a module z-score of 0.40 (CC 0.38). For the MDD group, the segmented module numbers of the cortex were 90.8 and 159.0 at gamma 0.45 and 0.90, respectively, with the module z-scores increasing from 0.56 (CC 0.50) to 0.76 (CC 0.64), see bottom subplot in Figure 2. The module number of the initial partition based on the Desikan-Killiany Atlas was 68, with an average voxel number of 131.1 (64mm^3/voxel) and a module z-score of 0.43 (CC 0.41). The MOSI-derived module z-scores outperformed their atlas-based counterparts and increased monotonously with higher gamma values, indicating that BOLD signals within a module became more homogeneous at finer resolutions, thereby confirming the validity of the MOSI scheme.

**Figure 2.**
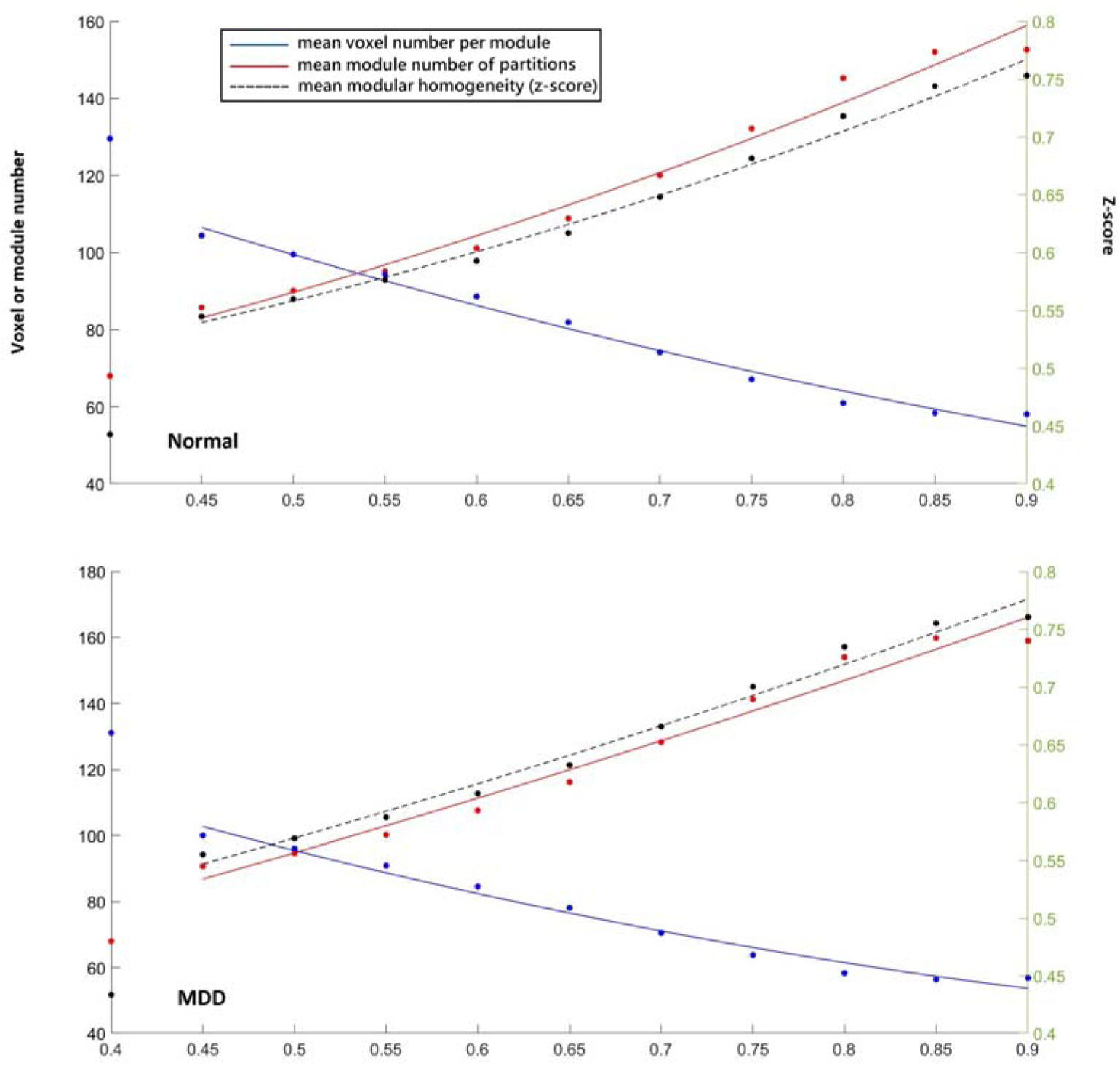
Scatter plots of three indices derived from CAN-BIND dataset, with X-axis gamma values and second-order polynomial regression fits. Top: Normal group, Bottom: MDD group. The origin (x = 0) represents the initial partition based on the Desikan-Killiany atlas. Red: mean module number of the partitions derived from different gamma values, referring to the left Y-axis. Blue: mean voxel number per module, referring to the left Y-axis. Black: mean module z-score (homogeneity within a module), referring to the right Y-axis.

For the between-group comparisons, independent t-tests revealed that the MDD group had a higher partition number and mean ALFF per voxel in the limbic compartment, but not in the frontoparietal compartment. Other indices, such as the mean voxel number per partition and total voxel number in each compartment, did not show significant statistical differences. Total ALFF showed a trend where the MDD group was higher than the Normal group in the limbic compartment, as summarized in Table 1. Previous reports indicated that MOSI’s results were comparable to those of existing network approaches at gamma values around 0.80–0.85 (Lee and Tramontano, 2021). Results at lower gamma values showed a similar pattern and are therefore included in the **Supplementary Materials** (Part I).

**Table 1.**
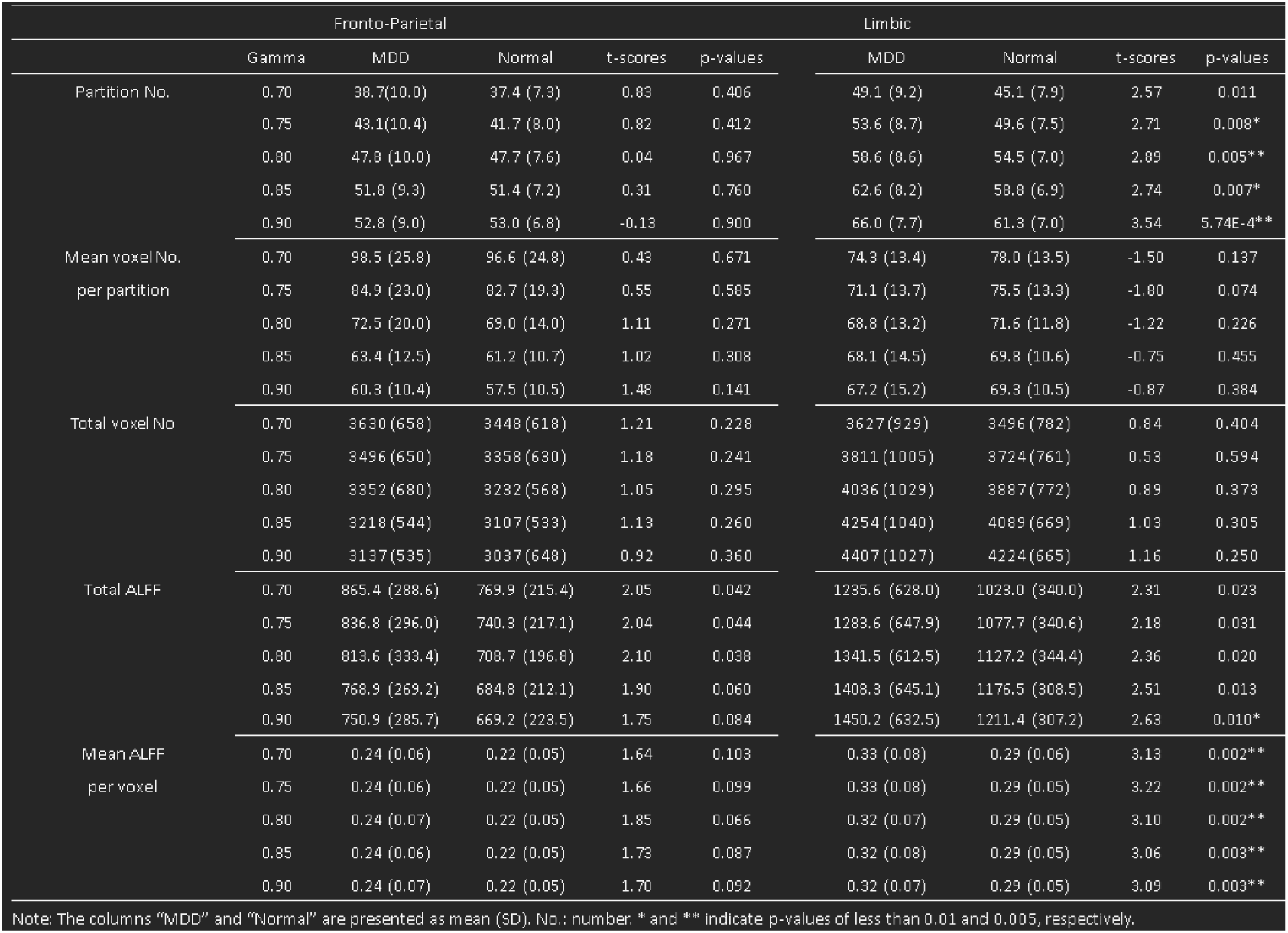
Statistics of the MOSI results (MDD - Normal)

### Inter-compartment power-connectivity relationship analyses

Consistent with previous reports, the correlation coefficients between nodal powers and nodal degrees were highly significant (p values < 1E-10) when combining the FP and limbic systems (class (3) of the Methods subsection: the relationship between power and connectivity). These coefficients ranged from -0.31 to -0.41 across all gamma values in both the MDD and Normal groups, with no significant differences between groups, see Table 2. Nevertheless, the main focus remains on the inter-compartment power-connectivity relationship, i.e., class (1) FP-Limbic and class (2) Limbic-FP, detailed below.

**Table 2.**
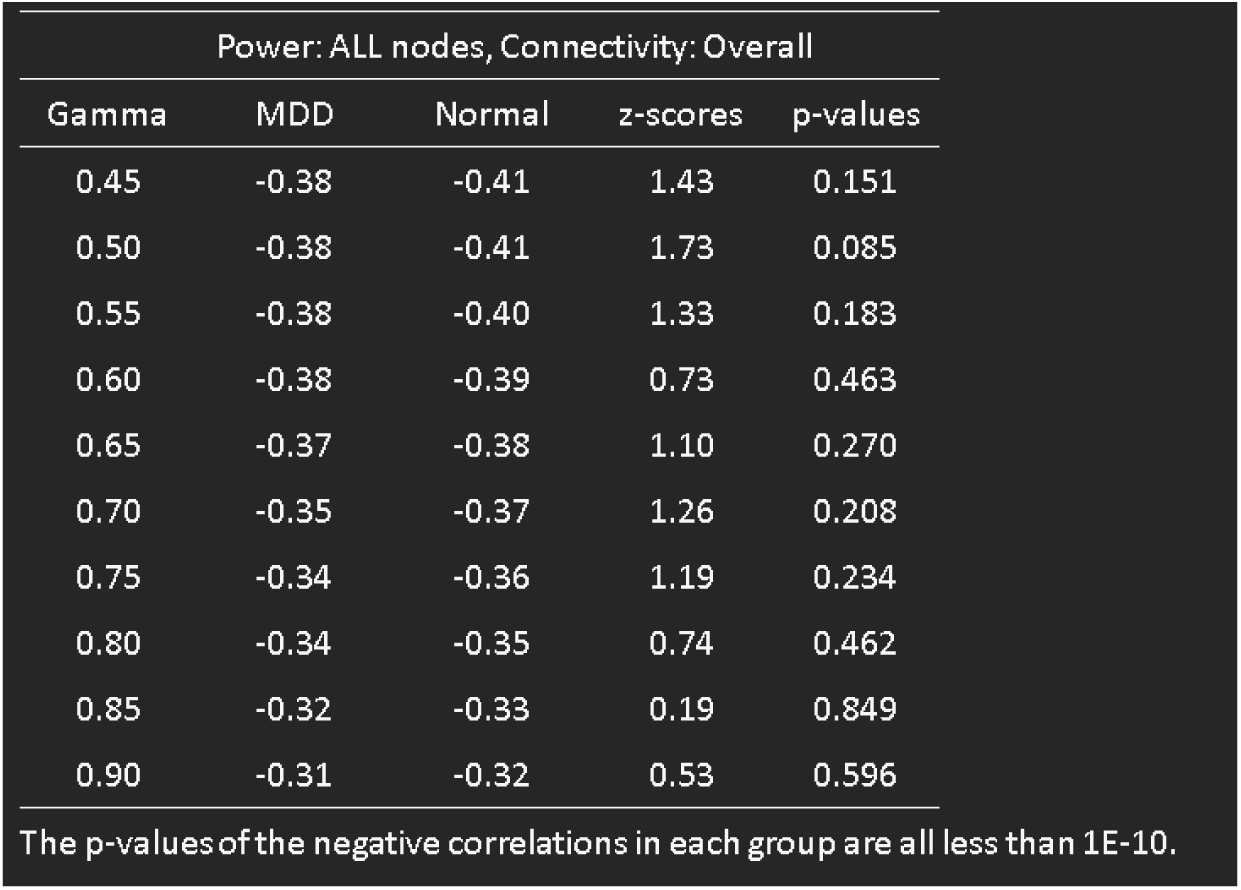
Correlation differences of overall power-connectivity relationship (MDD - Normal) Power: ALL nodes, Connectivity: Overall.

| Power: ALL nodes, Connectivity: Overall |  |  |  |  |
| --- | --- | --- | --- | --- |
| Gamma | MDD | Normal | z-scores | p-values |
| 0.45 | -0.38 | -0.41 | 1.43 | 0.151 |
| 0.50 | -0.38 | -0.41 | 1.73 | 0.085 |
| 0.55 | -0.38 | -0.40 | 1.33 | 0.183 |
| 0.60 | -0.38 | -0.39 | 0.73 | 0.463 |
| 0.65 | -0.37 | -0.38 | 1.10 | 0.270 |
| 0.70 | -0.35 | -0.37 | 1.26 | 0.208 |
| 0.75 | -0.34 | -0.36 | 1.19 | 0.234 |
| 0.80 | -0.34 | -0.35 | 0.74 | 0.462 |
| 0.85 | -0.32 | -0.33 | 0.19 | 0.849 |
| 0.90 | -0.31 | -0.32 | 0.53 | 0.596 |

The p-values of the negative correlations in each group are all less than 1E-10.

Class (1) FP-Limbic examined the relationship between mean FP power and the mean FC between the FP and limbic compartments, with the latter referred to as Limbic-to-FP FC for clarity, while acknowledging the bidirectional nature of FC. Similarly, class (2) Limbic-FP focused on the mean limbic power and mean inter-compartment FC, designated as FP-to-Limbic FC. Notably, under weighting strategy II, the mean FCs of Limbic-to-FP and FP-to-Limbic differ, requiring distinct naming for clarity. Moreover, the influence of connectivity between the FP and limbic systems on FP power must operate in a Limbic-to-FP direction. The same logic applies to FP-to-Limbic. Therefore, the deliberate abuse of notation is justified for the sake of clarity in this context. Significant differences in the inter-compartment power-connectivity relationship were observed between the MDD and Normal groups at gamma values 0.45 to 0.80 (p-values less than 0.01 or 0.005). Interestingly, opposite patterns are observed: For the FP nodal power and Limbic-to-FP connectivity, the MDD group showed a more negative correlation at all gamma values, whereas in the case of Limbic nodal power and FP-to-Limbic connectivity, the MDD group exhibited less negative correlation. These results together highlight that individuals with MDD display deviant FP-Limbic and Limbic-FP inter-compartmental interactions, illustrated in Figure 3 and summarized in Table 3.

**Figure 3.**
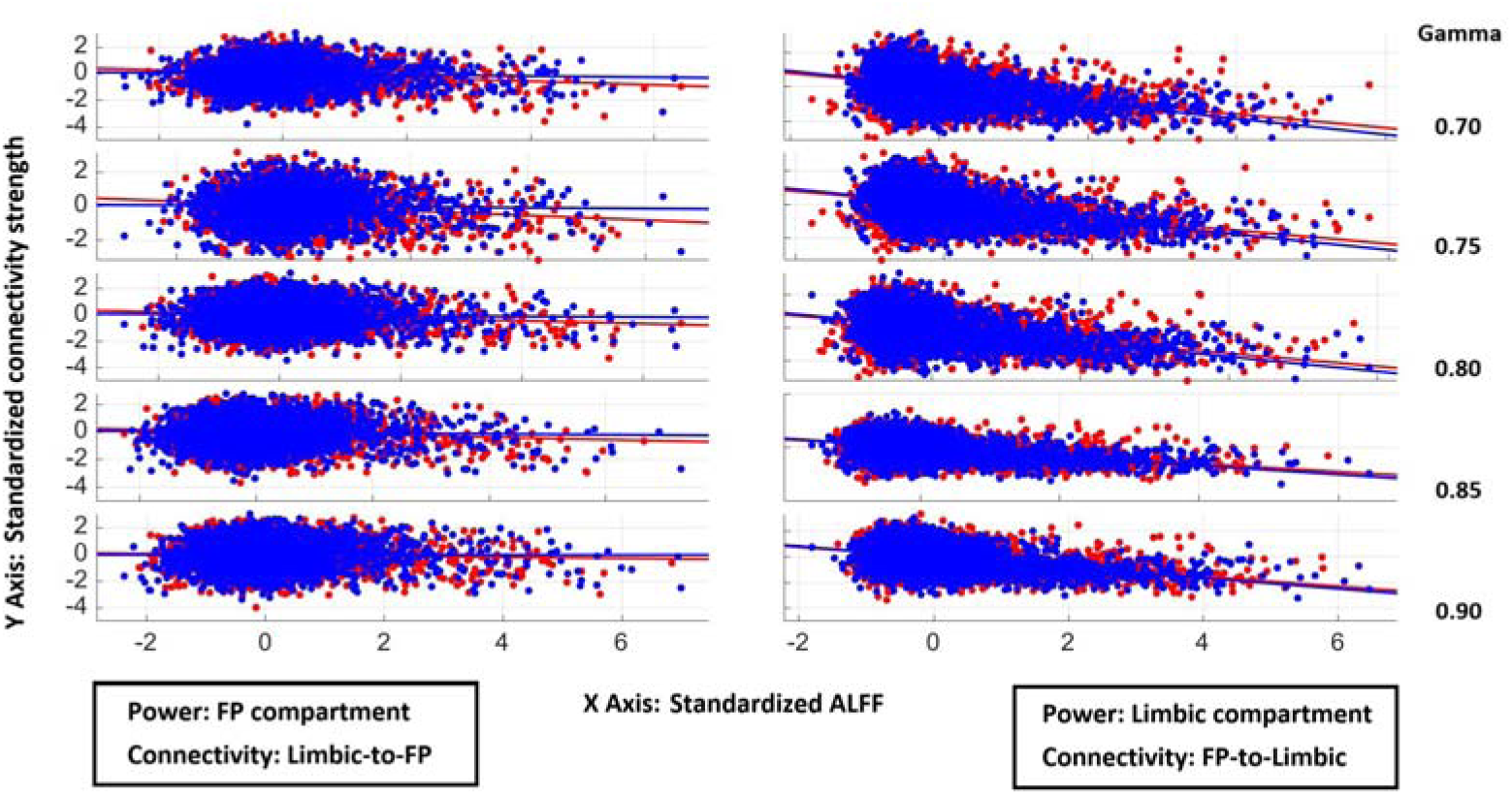
Scatter plots with regression lines illustrating the relationships between power and connectivity. Left column: FP powers and Limbic-to-FP connectivity strengths. Right column: Limbic powers and FP-to-Limbic connectivity strengths. The rows are sorted by five gamma values, ranging from 0.70 to 0.90. Blue indicates Normal, while Red represents MDD. Note that the blue lines are above the red lines in the left column and below the red lines in the right column, respectively.

**Table 3.** Correlation differences of inter-compartment power-connectivity relationship (MDD - Normal)

| Gamma | Power: FP nodes, Connectivity: FP-LM |  |  |  | Power: Limbic nodes, Connectivity: LM-FP |  |  |  |
| --- | --- | --- | --- | --- | --- | --- | --- | --- |
|  | MDD | Normal | z-scores | p-values | MDD | Normal | z-scores | p-values |
| 0.45 | -0.20 | -0.08 | -3.58 | 3.45E-4** | -0.42 | -0.49 | 2.89 | 0.004** |
| 0.50 | -0.19 | -0.09 | -3.23 | 0.001** | -0.42 | -0.50 | 3.05 | 0.002** |
| 0.55 | -0.18 | -0.08 | -3.00 | 0.003** | -0.43 | -0.50 | 2.84 | 0.004** |
| 0.60 | -0.18 | -0.06 | -3.97 | 7.26E-5** | -0.43 | -0.49 | 2.69 | 0.007* |
| 0.65 | -0.15 | -0.04 | -3.52 | 4.36E-4** | -0.42 | -0.47 | 2.61 | 0.009* |
| 0.70 | -0.14 | -0.04 | -3.27 | 0.001** | -0.40 | -0.46 | 2.98 | 0.003** |
| 0.75 | -0.14 | -0.02 | -4.06 | 4.98E-5** | -0.39 | -0.45 | 2.83 | 0.005** |
| 0.80 | -0.11 | -0.03 | -3.15 | 0.002** | -0.39 | -0.44 | 2.50 | 0.012 |
| 0.85 | -0.09 | -0.03 | -2.36 | 0.018 | -0.38 | -0.42 | 2.15 | 0.031 |
| 0.90 | -0.04 | -0.00 | -1.70 | 0.088 | -0.38 | -0.41 | 1.61 | 0.107 |

\* and \*\* indicate p-values of less than 0.01 and 0.005, respectively.

### QQ plot and ensuing analyses

A QQ plot was employed to assess the module size distribution in the MDD and Normal groups. It was observed that as the size increased beyond the top 15%, group-level differences emerged, see Figure 4. Opposite patterns were noticed again: MDD’s module sizes tended to be larger in the FP compartment (below the equal line in Figure 4) and lower in the limbic compartment (above the equal line in Figure 4), compared with Normal group. Given the observation, further exploratory analyses were conducted to compare the mean ALFFs and mean inter-compartment connectivity strengths between the largest 15% and the remaining 85% modules.

**Figure 4.**
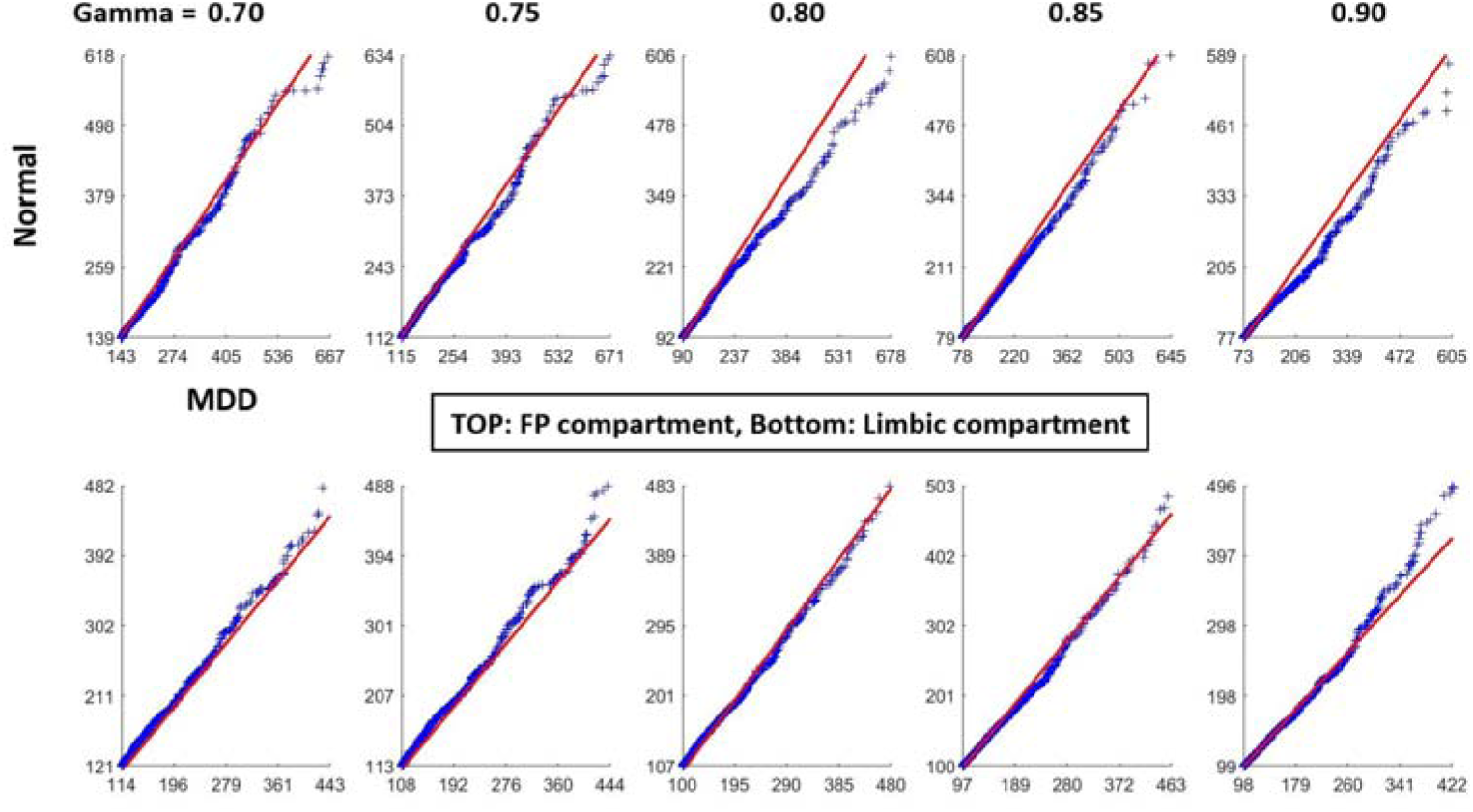
QQ plot of module size distribution (blue dots) for the largest 20% of modules in MDD and Normal groups. The sizes represented by both axes are equal along the red line.

For the Normal group, Table 4 demonstrates that larger modules are associated with lower mALFF, with significant results in the limbic compartment (p-values < 0.005 for Gamma 0.45 to 0.85) and a trend in the FP compartment (all t-scores are negative, with a non-parametric p-value < 0.001 based on binomial distribution). Regarding connectivity, both LM-to-FP and FP-to-LM FCs showed that the top 15% of modules had significantly greater connectivity strength compared to the remaining 85% across all gamma values (p-values < 0.005). The results suggest that larger modules play a more prominent role in inter-compartment interactions, while also supporting the notion that stronger global or inter-compartment FCs carry more inhibitory information (Lee and Tramontano, 2024).

**Table 4.**
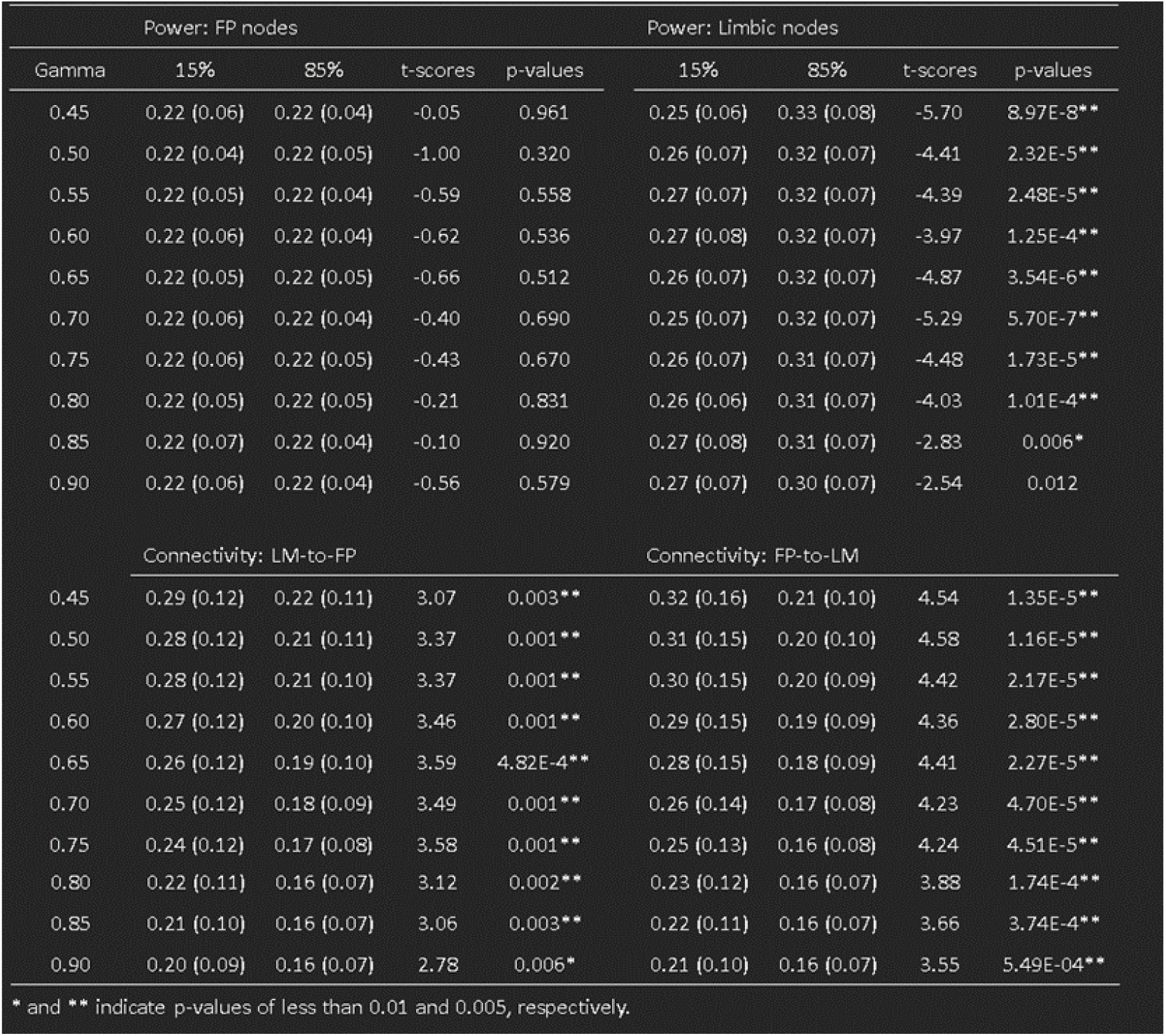
Comparisons of mean powerand connectivity strength between the 15% largest and the remaining 35% of modules(Normal.

| Gamma | Power: FP nodes |  |  |  | Power: Limbic nodes |  |  |  |
| --- | --- | --- | --- | --- | --- | --- | --- | --- |
|  | 15% | 85% | t-scores | p-values | 15% | 85% | t-scores | p-values |
| 0.45 | 0.22 (0.06) | 0.22 (0.04) | -0.05 | 0.961 | 0.25 (0.06) | 0.33 (0.08) | -5.70 | 8.97E-8** |
| 0.50 | 0.22 (0.04) | 0.22 (0.05) | -1.00 | 0.320 | 0.26 (0.07) | 0.32 (0.07) | -4.41 | 2.32E-5** |
| 0.55 | 0.22 (0.05) | 0.22 (0.04) | -0.59 | 0.558 | 0.27 (0.07) | 0.32 (0.07) | -4.39 | 2.48E-5** |
| 0.60 | 0.22 (0.06) | 0.22 (0.04) | -0.62 | 0.536 | 0.27 (0.08) | 0.32 (0.07) | -3.97 | 1.25E-4** |
| 0.65 | 0.22 (0.05) | 0.22 (0.05) | -0.66 | 0.512 | 0.26 (0.07) | 0.32 (0.07) | -4.87 | 3.54E-6** |
| 0.70 | 0.22 (0.06) | 0.22 (0.04) | -0.40 | 0.690 | 0.25 (0.07) | 0.32 (0.07) | -5.29 | 5.70E-7** |
| 0.75 | 0.22 (0.06) | 0.22 (0.05) | -0.43 | 0.670 | 0.26 (0.07) | 0.31 (0.07) | -4.48 | 1.73E-5** |
| 0.80 | 0.22 (0.05) | 0.22 (0.05) | -0.21 | 0.831 | 0.26 (0.06) | 0.31 (0.07) | -4.03 | 1.01E-4** |
| 0.85 | 0.22 (0.07) | 0.22 (0.04) | -0.10 | 0.920 | 0.27 (0.08) | 0.31 (0.07) | -2.83 | 0.006* |
| 0.90 | 0.22 (0.06) | 0.22 (0.04) | -0.56 | 0.579 | 0.27 (0.07) | 0.30 (0.07) | -2.54 | 0.012 |
| Gamma | Connectivity: LM-to-FP |  |  |  | Connectivity: FP-to-LM |  |  |  |
|  | 15% | 85% | t-scores | p-values | 15% | 85% | t-scores | p-values |
| 0.45 | 0.29 (0.12) | 0.22 (0.11) | 3.07 | 0.003** | 0.32 (0.16) | 0.21 (0.10) | 4.54 | 1.35E-5** |
| 0.50 | 0.28 (0.12) | 0.21 (0.11) | 3.37 | 0.001** | 0.31 (0.15) | 0.20 (0.10) | 4.58 | 1.16E-5** |
| 0.55 | 0.28 (0.12) | 0.21 (0.10) | 3.37 | 0.001** | 0.30 (0.15) | 0.20 (0.09) | 4.42 | 2.17E-5** |
| 0.60 | 0.27 (0.12) | 0.20 (0.10) | 3.46 | 0.001** | 0.29 (0.15) | 0.19 (0.09) | 4.36 | 2.80E-5** |
| 0.65 | 0.26 (0.12) | 0.19 (0.10) | 3.59 | 4.82E-4** | 0.28 (0.15) | 0.18 (0.09) | 4.41 | 2.27E-5** |
| 0.70 | 0.25 (0.12) | 0.18 (0.09) | 3.49 | 0.001** | 0.26 (0.14) | 0.17 (0.08) | 4.23 | 4.70E-5** |
| 0.75 | 0.24 (0.12) | 0.17 (0.08) | 3.58 | 0.001** | 0.25 (0.13) | 0.16 (0.08) | 4.24 | 4.51E-5** |
| 0.80 | 0.22 (0.11) | 0.16 (0.07) | 3.12 | 0.002** | 0.23 (0.12) | 0.16 (0.07) | 3.88 | 1.74E-4** |
| 0.85 | 0.21 (0.10) | 0.16 (0.07) | 3.06 | 0.003** | 0.22 (0.11) | 0.16 (0.07) | 3.66 | 3.74E-4** |
| 0.90 | 0.20 (0.09) | 0.16 (0.07) | 2.78 | 0.006* | 0.21 (0.10) | 0.16 (0.07) | 3.55 | 5.49E-4** |

\* and \*\* indicate p-values of less than 0.01 and 0.005, respectively.

For the MDD group, Table 5 summarizes the patterns in power and FC, noting that the latter is similar to, while the former differs from those in Table 4. The larger modules in both the FP and limbic compartments in the MDD group were associated with weaker inter-compartment interactions. However, this differential size effect was less pronounced in terms of power. It is noteworthy that since weighting strategy II was adopted, the FC is not influenced by the number of voxels in the node, but rather reflects the (input) connectivity strengths with the other compartment. The interaction of 2-way ANOVA was not significant (15% vs. 85% and MDD vs. Normal), and thus no further analyses were conducted.

**Table 5.** Com parisons of mean power and connectivity strength between the 15% largest and the remaining85% of modules(MDD)

| Gamma | Power: FP nodes |  |  |  | Power: Limbic nodes |  |  |  |
| --- | --- | --- | --- | --- | --- | --- | --- | --- |
|  | 15% | 85% | t-scores | p-values | 15% | 85% | t-scores | p-values |
| 0.45 | 0.24 (0.08) | 0.24 (0.06) | 0.55 | 0.585 | 0.30 (0.11) | 0.36 (0.10) | -3.03 | 0.003** |
| 0.50 | 0.24 (0.08) | 0.24 (0.06) | 0.15 | 0.883 | 0.30 (0.12) | 0.36 (0.11) | -2.65 | 0.009* |
| 0.55 | 0.24 (0.08) | 0.24 (0.06) | 0.09 | 0.930 | 0.31 (0.12) | 0.35 (0.09) | -2.01 | 0.047 |
| 0.60 | 0.24 (0.08) | 0.24 (0.06) | 0.03 | 0.973 | 0.31 (0.12) | 0.35 (0.09) | -2.40 | 0.018 |
| 0.65 | 0.24 (0.08) | 0.24 (0.07) | -0.31 | 0.757 | 0.30 (0.12) | 0.35 (0.09) | -2.43 | 0.017 |
| 0.70 | 0.24 (0.08) | 0.24 (0.05) | 0.00 | 0.998 | 0.31 (0.12) | 0.34 (0.09) | -1.74 | 0.085 |
| 0.75 | 0.24 (0.08) | 0.24 (0.05) | -0.02 | 0.986 | 0.31 (0.12) | 0.34 (0.08) | -1.87 | 0.064 |
| 0.80 | 0.24 (0.09) | 0.24 (0.06) | 0.45 | 0.651 | 0.31 (0.11) | 0.34 (0.08) | -1.38 | 0.169 |
| 0.85 | 0.24 (0.08) | 0.23 (0.06) | 0.08 | 0.937 | 0.32 (0.12) | 0.33 (0.07) | -0.21 | 0.835 |
| 0.90 | 0.24 (0.09) | 0.23 (0.05) | 0.30 | 0.763 | 0.33 (0.10) | 0.32 (0.08) | 0.32 | 0.748 |
| Gamma | Connectivity: LM-FP |  |  |  | Connectivity: FP-LM |  |  |  |
|  | 15% | 85% | t-scores | p-values | 15% | 85% | t-scores | p-values |
| 0.45 | 0.28 (0.16) | 0.21 (0.13) | 2.68 | 0.008* | 0.31 (0.18) | 0.20 (0.12) | 4.02 | 1.01E-4** |
| 0.50 | 0.28 (0.16) | 0.20 (0.12) | 2.94 | 0.004** | 0.31 (0.18) | 0.19 (0.12) | 4.17 | 5.92E-5** |
| 0.55 | 0.27 (0.15) | 0.20 (0.13) | 2.90 | 0.004** | 0.29 (0.17) | 0.19 (0.12) | 3.89 | 1.67E-4** |
| 0.60 | 0.26 (0.14) | 0.19 (0.12) | 2.88 | 0.005** | 0.28 (0.16) | 0.18 (0.11) | 4.10 | 7.60E-5** |
| 0.65 | 0.25 (0.14) | 0.18 (0.11) | 2.76 | 0.007* | 0.27 (0.17) | 0.17 (0.10) | 4.09 | 7.79E-5** |
| 0.70 | 0.24 (0.14) | 0.17 (0.11) | 2.93 | 0.004** | 0.25 (0.16) | 0.17 (0.10) | 3.70 | 3.30E-4** |
| 0.75 | 0.23 (0.13) | 0.16 (0.10) | 2.89 | 0.005** | 0.24 (0.15) | 0.16 (0.09) | 3.70 | 3.23E-4** |
| 0.80 | 0.21 (0.13) | 0.16 (0.09) | 2.71 | 0.008* | 0.22 (0.14) | 0.15 (0.09) | 3.02 | 0.003** |
| 0.85 | 0.20 (0.12) | 0.15 (0.08) | 2.58 | 0.011 | 0.20 (0.12) | 0.15 (0.08) | 2.88 | 0.005** |
| 0.90 | 0.20 (0.11) | 0.16 (0.09) | 2.40 | 0.018 | 0.20 (0.12) | 0.15 (0.08) | 2.79 | 0.006* |

\* and \*\* indicate p-values of less than 0.01 and 0.005, respectively.

## Discussion

The neural mechanisms underlying MDD have been a challenging issue in brain science. Extensive research has been integrated into the limbic-cortical dysregulation model (Mayberg, 1997), which provides an insightful framework. However, findings from independent studies of MDD are widely distributed across the PFC, parietal cortex, and limbic regions. The compromised circuits involve, but are not limited to, emotion regulation, emotion processing, reward, saliency, and executive control (Brand et al., 2015; Phillips et al., 2015; Pimontel et al., 2016). Moreover, abnormalities in just one or a few circuits cannot fully explain the multifaceted symptomatology of MDD. Thus, an effort has been made to expand the limbic-cortical dysregulation model to encompass the FP and limbic compartments, particularly emphasizing their imbalanced reciprocal suppression (briefly summarized in the section: *Potential mechanism mediating the FP and limbic imbalances*) (Lee, 2016; Lee and Xue, 2018b). Notably, both the compartments are well-acknowledged neural entities (Mega et al., 1997; Lee et al., 2014; Lee and Xue, 2018a), and the compartment-level model provides broader explanatory power than models focused on specific circuits, as it may better account for the subtypes of MDD and heterogeneity in symptomatology through compensatory changes and neural reorganization/plasticity (Lee et al., 2011; Boessen et al., 2012; Lee, 2016; Drysdale et al., 2017; Lee and Xue, 2018b). Despite its simplicity and biological plausibility, two major obstacles hinder the empirical verification of this extended model. First, an individualized and physiologically constrained functional parcellation of cortical signals (i.e., FCP) is required to enable whole-brain and compartment-level analyses. Second, due to the lower sampling rate inherent in fMRI compared to electroencephalography (EEG), mathematical models like Granger causality or isolated effective coherence (Barnett and Seth, 2014; Pascual-Marqui et al., 2014) are not feasible for investigating inter-compartmental causal interactions, which are expected to occur within tens to hundreds of milliseconds (Lee et al., 2023). These two issues have been addressed by MOSI (Lee and Tramontano, 2021), which facilitates FCP, and by the recent discovery of a robust negative relationship between nodal degree and nodal power (Lee and Tramontano, 2024), enabling the study of inhibitory interaction using resting state fMRI (rsfMRI)—specially, nodal degree may causally influence nodal power.

### The neuropathology of MDD unveiled through MOSI-facilitated compartment analyses

Statistical analyses revealed that MDD is associated with a higher partition number and greater mean ALFF per voxel in the limbic compartment. These findings not only align with the observed limbic hyperactivity (elevated mALFF) but also indicate that limbic neural activities become more heterogeneous and differentiated (increased module number). It is imperative to note that the insight provided this study is at the compartmental level, in contrast to the sporadic brain regions reported by conventional fMRI analyses, which lack consistency and are sometimes hard to explain. For example, both increased and decreased ALFFs have been reported in the limbic system, even within a single study (Jiao et al., 2011; Hou et al., 2022).

Reproducibility has been a persistent challenge in neuroimaging research, particularly in neuropsychiatric studies (Holiga et al., 2018). This research may offer novel insights into addressing this critical issue. Compartment-level exploration occurs at a broader or coarser scale, with the abnormalities likely triggering compensatory and plasticity changes at finer scales to counteract dysfunction. Evidence suggests that neural plasticity and reorganization are highly individualized processes (Poldrack, 2000; Kanai and Rees, 2011; Kolb and Gibb, 2014). Consequently, neural metrics across different scales may exhibit varying degrees of replicability, with the appropriate resolution expected to yield the highest consistency. An outstanding feature of MOSI is its multi-resolution investigation. By adjusting the resolutions through different Gamma values, the optimal resolution that reveals the key abnormality in MDD can be identified, which will be discussed later. Another key advantage of MOSI is its full consideration of individuality. As shown in Table 1, the variance in module number, module size and compartment size underscores MOSI’s suitability for imaging studies characterized by heterogeneity, such as in the case of MDD. This stands in sharp contrast to fixed partitions informed by anatomical atlases (Lee et al., 2014; Lee and Xue, 2017a). The variability in partition size is effectively managed through a weighting strategy, laying the groundwork for subsequent network analyses.

In the FP compartment, however, reduced ALFF as an indicator of hypoactivity was expected but not present. MDD even demonstrated a mild trend of increased FP mALFF across all resolutions (Table 1 and Part I in **Supplementary Material**). It is noteworthy that previous research has also failed to demonstrate increased ALFF in the prefrontal cortex of individuals with MDD (Hou et al., 2022). Some studies even indicated that MDD was associated with enhanced regional power (ALFF) in the prefrontal cortex (Jiao et al., 2011). It is reasonable to assume the engagement of compensatory mechanisms to alleviate FP hypoactivity. This conjecture is particularly applicable to the FP compartment, given its diverse cortical and subcortical inputs (Alexander et al., 1986; Shipp, 2004; Arnsten, 2013), which extend well beyond the limbic system. In this context, a trend of increased ALFF as a compensation in the FP conversely suggests its hypoactivity. Accordingly, it is inappropriate to oversimplify the concepts of hyperactivity and hypoactivity as merely increased or decreased regional power (ALFF). Instead, they may be better understood as relative concepts, for example, reflected in the altered interactions between the FP and limbic compartments, as discussed in the next section.

A QQ plot assessed module size distribution in the MDD and Normal groups, prompting a comparison of mean ALFFs and inter-compartment connectivity strengths between the largest 15% and the remaining 85% of modules. Larger modules were associated with lower mALFF and greater connectivity strength, suggesting a more prominent role in inter-compartment interactions. These findings reinforce the argument that nodal degree in rsfMRI carries more information about inhibitory coupling. However, the two-way ANOVA interaction analyses of mALFF and connectivity strengths were both negative, and for the MDD group, the results mirrored those in Table 1 and Part I of the supplementary material. Nevertheless, there appears to be an interesting trend in mALFF for the larger limbic modules in MDD: compared to their 85% counterparts, the 15% largest modules exhibited lower mALFF in MDD as expected, but the decrease was less pronounced than in the Normal group. This finding implies a relative limbic hyperactivity in the larger modules and suggests their possible role in maintaining the depressive state; please see Part III of the supplementary material for details.

### The insight derived from inter-compartmental power-connectivity analyses

The power-FC relationship within a large-scale network may serve as a surrogate for studying causal and inhibitory coupling in resting-state fMRI. The former indicates that nodal degree influences nodal power, not vice versa, while the latter reflects a negative correlation. This discovery was inspired by my previous report (nodal degree derived from SC vs nodal power ALFF) and an anecdotal observation that in-house software frequently revealed abnormally increased regional power in patients characterized by diffuse hypo-connections (Lee and Xue, 2017b; Lee and Tramontano, 2024).

For both the MDD and Normal groups, combining the FP and limbic compartments as a whole revealed robust negative correlation between nodal degree and nodal power, replicating a previous report (Table 2) (Lee and Tramontano, 2024). However, when focusing on the FCs between the FP and limbic compartments and investigating their influence on nodal power, interesting patterns unfolded. Firstly, the negative relationship is asymmetric, with regulation from the FP compartment to the LM compartment being much stronger than from the limbic to the FP compartment. This asymmetry aligns with literature on emotion regulation, cognitive therapy, and the effects of transcranial magnetic stimulation over the prefrontal cortex in alleviating depression severity (Speer et al., 2000; Taylor and Liberzon, 2007; McRae et al., 2012; Lee and Xue, 2018d). Throughout evolution, the prefrontal cortex has expanded significantly, particularly in humans and higher primates, enhancing its role in regulating behaviors and emotions. This development has led to greater top-down control over the limbic system, especially in modulating emotional responses from regions like the amygdala (Miller and Cohen, 2001; Fuster, 2002; Phillips et al., 2008). As a result, the influence of the prefrontal cortex on the limbic system has gradually surpassed the limbic system’s reciprocal suppressive effects on the prefrontal cortex (Liberzon et al., 2003; Phan et al., 2004), reflected in the asymmetry of the nodal power and degree relationships.

Secondly, the two inter-compartmental relationships in the MDD group deviated in opposite directions compared to the Normal group. Specifically, the MDD group’s slopes of FP nodal power and nodal degree (i.e., FP power vs LM-to-FP connectivity) became steeper, while those of limbic nodal power and nodal degree (i.e., limbic power vs FP-to-LM connectivity) became flatter. This can be observed in the regression lines representing the relationship between power and connectivity strength in the scatter plots shown in Figure 3, as well as in the significant statistical comparisons summarized in Table 3. The results suggest that the MDD group is characterized by more efficient suppression from the limbic to frontal compartments and less efficient suppression in the reverse direction.

This intriguing pattern prompts a reappraisal of the implications of FP hypoactivity and limbic hyperactivity in MDD. As discussed in the previous section, the absolute magnitude of power is not an ideal measure for representing the concepts of hypoactivity and hyperactivity; rather, these should be assessed in a relative manner. The relativism revealed by the nodal-power vs nodal-degree analyses indicates an imbalanced interaction between the FP and limbic compartments. The hypoactive FP compartment may arise from and/or be maintained by abnormally stronger suppression from the limbic counterpart, while the hyperactive limbic compartment may originate from and/or be maintained by abnormally weaker suppression from the FP counterpart. This MDD theory of inter-compartmental network imbalance is thus empirically verified, expanding the insightful limbic-cortical dysregulation model proposed by Mayberg, and shedding light on a paramount issue of MDD: its genesis and maintenance (see the section on *potential mechanism mediating the FP and limbic imbalances*). The consequences of imbalanced reciprocal suppression can be readily applied to activation design experiments, which are beyond the scope of this article. The neural metrics from activation designs in MDD exhibit a stronger alignment with the characterization of hypoactive FP and hyperactive limbic systems, as noted in the summary of previous research (Lee and Xue, 2018b).

One merit of the multi-resolution approach lies in its potential to provide the appropriate scale(s) for unveiling the neuropathology of various neuropsychiatric conditions (or other scientific issues). The analyses above revealed that the most informative Gamma values for partition size, mALFF, and power-connectivity correlation are 0.55 to 0.90, 0.45 to 0.90, and 0.45 to 0.75, respectively. Taken together, Gamma values ranging from 0.55 to 0.75 seem to offer suitable resolutions for exploring the neuropathology of MDD, which is lower than the recommended range for reproducing canonical networks (i.e., Gamma values of 0.80 to 0.85 (Lee and Tramontano, 2021)). Such resolution-dependent findings are conceptually linked to the various scales inherent in the hierarchical organization of brain tissues (Meunier et al., 2009) and warrants further research to investigate.

### Potential mechanism mediating the FP and limbic imbalances

Humans universally experience switching between exteroception and interoception, a process supported by two major networks: the FP mantle and the limbic system (Lee et al., 2014). I propose that a reciprocal suppressive mechanism (RSM) must exist to facilitate fluid switching between these two states/compartments. Further, the RSM must be maintained in a dynamic equilibrium. If this balance is disrupted, a vicious cycle may occur and the consequences is catastrophic, potentially leading to MDD, as illustrated in Figure 5. Various medical conditions may disrupt the inter-compartment balance. For instance, prolonged stress can simultaneously enhance limbic activity and attenuate FP activity (Diorio et al., 1993; Arnsten, 2009). Additionally, vascular insufficiency in the white matter may impair FP functioning (O’Sullivan et al., 2001). When the cortico-limbic imbalance reaches a critical threshold, limbic hyperactivity (relative to the FP) may amplify its inhibitory effect on the FP compartment, reducing the FP’s counter-regulatory influence on the limbic system. This imbalance creates a vicious cycle, stabilizing into the hyper-limbic/hypo-FP state characteristic of MDD. For a detailed formulation and methodology of the RSM theory of MDD, please refer to (Lee, 2016; Lee and Xue, 2018b; Lee, 2024).

**Figure 5.**
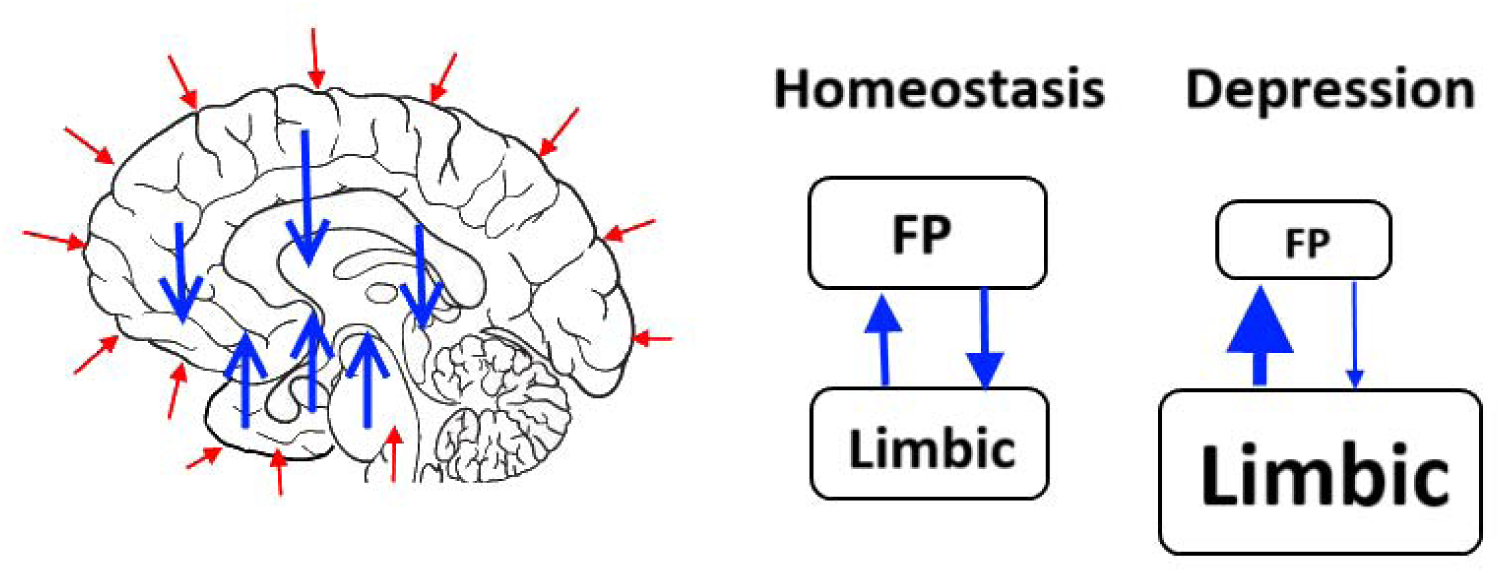
Highlighted interacting patterns of two networks related to MDD: reciprocal suppression between the FP and limbic compartments. Left: Short red arrows indicate various excitatory inputs to both compartments of the brain. Blue arrows represent mutual suppression as an interaction mechanism between the two compartments. Right: Under homeostasis, balance is achieved, with the sizes of the dorsal and ventral compartments (abstractly representing metabolic or activity levels) approximately equal and allowing for mild perturbations. When this balance is disrupted to a breaking point, the limbic compartment enlarges (conceptually), increasing its negative influence on the dorsal compartment; conversely, the dorsal compartment shrinks, reducing its impact on the ventral limbic system. The overall consequence evolves into a state/attractor of FP hypoactivity and limbic hyperactivity, characteristic of MDD.

## Conclusion

This study provides empirical validation of the framework of imbalanced reciprocal suppression between the FP and limbic compartments, designed to address the central neuropathology of MDD. This inter-compartmental imbalance framework, mediated by abnormal RSM, extends and refines the cortico-limbic dysregulation model, explaining the observed hypoactivity in the FP and hyperactivity in the limbic system. Additionally, the resulting neural compensatory changes and reorganization through plasticity may account for individual differences in symptomatology and subtypes of MDD, which can be empirically examined in future studies. Furthermore, this study highlights an important caveat: the neural metric ALFF should not be reported in isolation; it is essential to consider nodal degree due to their strong negative correlation.

## Data Availability

All data produced in the present work are contained in the manuscript

## Authors Contributions

This report is authored by a single individual.

## Declarations

### Ethical Approval

This study was approved by Pearl IRB (ID 2023-0133; exempt category).

### Availability of data and materials

This research analyzed the databank from a publicly released dataset Canadian Biomarker Integration Network in Depression (CAN-BIND).

## Compliance with ethical standards

The author carried out no animal or human studies for this article. No conflicts of interest to declare.

## References

Aihara M, Ida I, Yuuki N, Oshima A, Kumano H, Takahashi K, Fukuda M, Oriuchi N, Endo K, Matsuda H, Mikuni M (2007) Hpa axis dysfunction in unmedicated major depressive disorder and its normalization by pharmacotherapy correlates with alteration of neural activity in prefrontal cortex and limbic/paralimbic regions. Psychiatry Res 155:245–256.

Alexander GE, DeLong MR, Strick PL (1986) Parallel organization of functionally segregated circuits linking basal ganglia and cortex. Annu Rev Neurosci 9:357–381.

American Psychiatric Association (2013) Diagnostic and statistical manual of mental disorders (DSM-5®): American Psychiatric Pub.

Amunts K, Schleicher A, Burgel U, Mohlberg H, Uylings HB, Zilles K (1999) Broca’s region revisited: cytoarchitecture and intersubject variability. J Comp Neurol 412:319–341.

Arnsten AF (2009) Stress signalling pathways that impair prefrontal cortex structure and function. Nature reviews neuroscience 10:410–422.

Arnsten AF (2013) The neurobiology of thought: the groundbreaking discoveries of Patricia Goldman-Rakic 1937–2003. Cerebral Cortex 23:2269–2281.

Bagherzadeh-Azbari S, Khazaie H, Zarei M, Spiegelhalder K, Walter M, Leerssen J, Van Someren EJ, Sepehry AA, Tahmasian M (2019) Neuroimaging insights into the link between depression and insomnia: a systematic review. J Affect Disord 258:133–143.

Barnett L, Seth AK (2014) The MVGC multivariate Granger causality toolbox: a new approach to Granger-causal inference. J Neurosci Methods 223:50–68.

Boessen R, Groenwold RH, Knol MJ, Grobbee DE, Roes KC (2012) Classifying responders and non-responders; does it help when there is evidence of differentially responding patient groups? J Psychiatr Res 46:1169–1173.

Brand SJ, Moller M, Harvey BH (2015) A Review of Biomarkers in Mood and Psychotic Disorders: A Dissection of Clinical vs. Preclinical Correlates. Current Neuropharmacology 13:324–368.

Brière FN, Rohde P, Seeley JR, Klein D, Lewinsohn PM (2014) Comorbidity between major depression and alcohol use disorder from adolescence to adulthood. Compr Psychiatry 55:526–533.

Busatto GF (2013) Structural and functional neuroimaging studies in major depressive disorder with psychotic features: A critical review. Schizophr Bull 39:776–786.

Chen TJ, Yu YW, Chen MC, Wang SY, Tsai SJ, Lee TW (2005) Serotonin dysfunction and suicide attempts in major depressives: An auditory event-related potential study. Neuropsychobiology 52:28–36.

Chialvo DR (2010) Emergent complex neural dynamics. Nature physics 6:744–750.

Cox RW (1996) AFNI: software for analysis and visualization of functional magnetic resonance neuroimages. Comput Biomed Res 29:162–173.

Culang-Reinlieb ME, Johnert LC, Brickman AM, Steffens DC, Garcon E, Sneed JR (2010) Mri-defined vascular depression: A review of the construct. Int J Geriatr Psychiatry.

Dale AM, Fischl B, Sereno MI (1999) Cortical surface-based analysis. I. Segmentation and surface reconstruction. Neuroimage 9:179–194.

Damoiseaux JS, Rombouts SA, Barkhof F, Scheltens P, Stam CJ, Smith SM, Beckmann CF (2006) Consistent resting-state networks across healthy subjects. Proc Natl Acad Sci U S A 103:13848–13853.

Desikan RS, Segonne F, Fischl B, Quinn BT, Dickerson BC, Blacker D, Buckner RL, Dale AM, Maguire RP, Hyman BT, Albert MS, Killiany RJ (2006) An automated labeling system for subdividing the human cerebral cortex on MRI scans into gyral based regions of interest. Neuroimage 31:968–980.

Diorio D, Viau V, Meaney MJ (1993) The role of the medial prefrontal cortex (cingulate gyrus) in the regulation of hypothalamic-pituitary-adrenal responses to stress. J Neurosci 13:3839–3847.

Disner SG, Beevers CG, Haigh EA, Beck AT (2011) Neural mechanisms of the cognitive model of depression. Nat Rev Neurosci 12:467–477.

Downar J, Geraci J, Salomons TV, Dunlop K, Wheeler S, McAndrews MP, Bakker N, Blumberger DM, Daskalakis ZJ, Kennedy SH, Flint AJ, Giacobbe P (2014) Anhedonia and reward-circuit connectivity distinguish nonresponders from responders to dorsomedial prefrontal repetitive transcranial magnetic stimulation in major depression. Biol Psychiatry 76:176–185.

Drevets WC (1999) Prefrontal cortical-amygdalar metabolism in major depression. Ann N Y Acad Sci 877:614–637.

Drevets WC, Bogers W, Raichle ME (2002) Functional anatomical correlates of antidepressant drug treatment assessed using pet measures of regional glucose metabolism. Eur Neuropsychopharmacol 12:527–544.

Drevets WC, Price JL, Furey ML (2008) Brain structural and functional abnormalities in mood disorders: Implications for neurocircuitry models of depression. Brain Struct Funct 213:93–118.

Drysdale AT et al. (2017) Resting-state connectivity biomarkers define neurophysiological subtypes of depression. Nat Med 23:28–38.

Elliott R, Lythe K, Lee R, McKie S, Juhasz G, Thomas EJ, Downey D, Deakin JF, Anderson IM (2012) Reduced medial prefrontal responses to social interaction images in remitted depression. Arch Gen Psychiatry 69:37–45.

Ferrari AJ, Charlson FJ, Norman RE, Flaxman AD, Patten SB, Vos T, Whiteford HA (2013) The epidemiological modelling of major depressive disorder: application for the Global Burden of Disease Study 2010. PLoS ONE 8:e69637.

Fischl B, Sereno MI, Dale AM (1999) Cortical surface-based analysis. II: Inflation, flattening, and a surface-based coordinate system. Neuroimage 9:195–207.

Freeman WJ, Chang HJ, Burke BC, Rose PA, Badler J (1997) Taming chaos: stabilization of aperiodic attractors by noise [olfactory system model]. IEEE Transactions on Circuits and Systems I: Fundamental Theory and Applications 44:989–996.

Fuster JM (2002) Frontal lobe and cognitive development. J Neurocytol 31:373–385.

Heinzel A, Grimm S, Beck J, Schuepbach D, Hell D, Boesiger P, Boeker H, Northoff G (2009) Segregated neural representation of psychological and somatic-vegetative symptoms in severe major depression. Neurosci Lett 456:49–53.

Henry ME, Kaufman MJ, Hennen J, Michelson D, Schmidt ME, Stoddard E, Vukovic AJ, Barreira PJ, Cohen BM, Renshaw PF (2003) Cerebral blood volume and clinical changes on the third day of placebo substitution for ssri treatment. Biol Psychiatry 53:100–105.

Holiga Š, Sambataro F, Luzy C, Greig G, Sarkar N, Renken RJ, Marsman JC, Schobel SA, Bertolino A, Dukart J (2018) Test-retest reliability of task-based and resting-state blood oxygen level dependence and cerebral blood flow measures. PLoS One 13:e0206583.

Holthoff VA, Beuthien-Baumann B, Zundorf G, Triemer A, Ludecke S, Winiecki P, Koch R, Fuchtner F, Herholz K (2004) Changes in brain metabolism associated with remission in unipolar major depression. Acta Psychiatr Scand 110:184–194.

Honey CJ, Sporns O, Cammoun L, Gigandet X, Thiran JP, Meuli R, Hagmann P (2009) Predicting human resting-state functional connectivity from structural connectivity. Proc Natl Acad Sci U S A 106:2035–2040.

Hou X, Mei B, Wang F, Guo H, Li S, Wu G, Zang C, Cao B (2022) Neural activity in adults with major depressive disorder differs from that in healthy individuals: A resting-state functional magnetic resonance imaging study. Front Psychiatry 13:1028518.

Hwang JP, Lee TW, Tsai SJ, Chen TJ, Yang CH, Lirng JF, Tsai CF (2010) Cortical and subcortical abnormalities in late-onset depression with history of suicide attempts investigated with mri and voxel-based morphometry. J Geriatr Psychiatry Neurol 23:171–184.

Jiao Q, Ding J, Lu G, Su L, Zhang Z, Wang Z, Zhong Y, Li K, Ding M, Liu Y (2011) Increased activity imbalance in fronto-subcortical circuits in adolescents with major depression. PLoS ONE 6:e25159.

Jo HJ, Saad ZS, Simmons WK, Milbury LA, Cox RW (2010) Mapping sources of correlation in resting state FMRI, with artifact detection and removal. Neuroimage 52:571–582.

Kanai R, Rees G (2011) The structural basis of inter-individual differences in human behaviour and cognition. Nat Rev Neurosci 12:231–242.

Kennedy SH, Evans KR, Kruger S, Mayberg HS, Meyer JH, McCann S, Arifuzzman AI, Houle S, Vaccarino FJ (2001) Changes in regional brain glucose metabolism measured with positron emission tomography after paroxetine treatment of major depression. Am J Psychiatry 158:899–905.

Kessler RC, Gruber M, Hettema JM, Hwang I, Sampson N, Yonkers KA (2008) Co-morbid major depression and generalized anxiety disorders in the national comorbidity survey follow-up. Psychol Med 38:365–374.

Kolb B, Gibb R (2014) Searching for the principles of brain plasticity and behavior. Cortex 58:251–260.

Lam RW, Milev R, Rotzinger S, Andreazza AC, Blier P, Brenner C, Daskalakis ZJ, Dharsee M, Downar J, Evans KR (2016) Discovering biomarkers for antidepressant response: protocol from the Canadian biomarker integration network in depression (CAN-BIND) and clinical characteristics of the first patient cohort. BMC psychiatry 16:1–13.

Lee T-W (2016) Network balance and its relevance to affective disorders: Dialectic neuroscience: Pronoun.

Lee T-W, Xue S-W (2017a) Examination of the validity of the atlas-informed approach to functional parcellation: a resting functional MRI study. Neuroreport 28:649–653.

Lee TW (2024) A novel form of dialectics informed by psychiatry and brain science: centralized dialectics.

Lee TW, Xue SW (2017b) Linking graph features of anatomical architecture to regional brain activity: A multi-modal MRI study. Neurosci Lett 651:123–127.

Lee TW, Xue SW (2018a) Functional connectivity maps based on hippocampal and thalamic dynamics may account for the default-mode network. Eur J Neurosci 47:388–398.

Lee TW, Xue SW (2018b) Extended cortico-limbic dysregulation model of major depressive disorder: a demonstration of the application of an analysis-synthesis framework to explore psychopathology. International Journal of Psychology Research 11:247–297.

Lee TW, Xue SW (2018c) Revisiting the Functional and Structural Connectivity of Large-Scale Cortical Networks. Brain Connect 8:129–138.

Lee TW, Xue SW (2018d) Does emotion regulation engage the same neural circuit as working memory? A meta-analytical comparison between cognitive reappraisal of negative emotion and 2-back working memory task. PLoS ONE 13:e0203753.

Lee TW, Tramontano G (2021) Automatic parcellation of resting-state cortical dynamics by iterative community detection and similarity measurements. AIMS neuroscience 8:526–542.

Lee TW, Tramontano G (2024) Negative relationship between inter-regional interaction and regional power: a resting fMRI study medRxiv 2023101323297027.

Lee TW, Northoff G, Wu YT (2014) Resting network is composed of more than one neural pattern: An fMRI study. Neuroscience 274:198–208.

Lee TW, Tramontano G, Hinrichs C (2023) Concordant dynamic changes of global network properties in the frontoparietal and limbic compartments: An EEG study. Biosystems 235:105101.

Lee TW, Yu YW, Chen MC, Chen TJ (2011) Cortical mechanisms of the symptomatology in major depressive disorder: A resting eeg study. J Affect Disord 131:243–250.

Lee TW, Liu HL, Wai YY, Ko HJ, Lee SH (2013) Abnormal neural activity in partially remitted late-onset depression: An fmri study of one-back working memory task. Psychiatry Res 213:133–141.

Liberzon I, Phan KL, Decker LR, Taylor SF (2003) Extended amygdala and emotional salience: A pet activation study of positive and negative affect. Neuropsychopharmacology 28:726–733.

Liotti M, Mayberg HS, McGinnis S, Brannan SL, Jerabek P (2002) Unmasking disease-specific cerebral blood flow abnormalities: Mood challenge in patients with remitted unipolar depression. Am J Psychiatry 159:1830–1840.

Mayberg HS (1997) Limbic-cortical dysregulation: A proposed model of depression. J Neuropsychiatry Clin Neurosci 9:471–481.

Mayberg HS (2003) Modulating dysfunctional limbic-cortical circuits in depression: Towards development of brain-based algorithms for diagnosis and optimised treatment. Br Med Bull 65:193–207.

Mayberg HS, Brannan SK, Tekell JL, Silva JA, Mahurin RK, McGinnis S, Jerabek PA (2000) Regional metabolic effects of fluoxetine in major depression: Serial changes and relationship to clinical response. Biol Psychiatry 48:830–843.

Mayberg HS, Lozano AM, Voon V, McNeely HE, Seminowicz D, Hamani C, Schwalb JM, Kennedy SH (2005) Deep brain stimulation for treatment-resistant depression. Neuron 45:651–660.

McRae K, Gross JJ, Weber J, Robertson ER, Sokol-Hessner P, Ray RD, Gabrieli JD, Ochsner KN (2012) The development of emotion regulation: An fmri study of cognitive reappraisal in children, adolescents and young adults. Soc Cogn Affect Neurosci 7:11–22.

Mega MS, Cummings JL, Salloway S, Malloy P (1997) The limbic system: An anatomic, phylogenetic, and clinical perspective. J Neuropsychiatry Clin Neurosci 9:315–330.

Meunier D, Lambiotte R, Fornito A, Ersche KD, Bullmore ET (2009) Hierarchical modularity in human brain functional networks. Front Neuroinform 3:37.

Miller EK, Cohen JD (2001) An integrative theory of prefrontal cortex function. Annu Rev Neurosci 24:167–202.

O’Sullivan M, Jones DK, Summers P, Morris R, Williams S, Markus H (2001) Evidence for cortical “disconnection” as a mechanism of age-related cognitive decline. Neurology 57:632–638.

Oquendo M, Brent DA, Birmaher B, Greenhill L, Kolko D, Stanley B, Zelazny J, Burke AK, Firinciogullari S, Ellis SP (2005) Posttraumatic stress disorder comorbid with major depression: factors mediating the association with suicidal behavior. Am J Psychiatry 162:560–566.

Pascual-Marqui R, Biscay R, Bosch-Bayard J, Lehmann D, Kochi K, Yamada N, Kinoshita T, Sadato N (2014) Isolated effective coherence (iCoh): causal information flow excluding indirect paths. arXiv preprint arXiv:14024887.

Phan KL, Taylor SF, Welsh RC, Ho S-H, Britton JC, Liberzon I (2004) Neural correlates of individual ratings of emotional salience: A trial-related fmri study. Neuroimage 21:768–780.

Phillips ML, Ladouceur CD, Drevets WC (2008) A neural model of voluntary and automatic emotion regulation: Implications for understanding the pathophysiology and neurodevelopment of bipolar disorder. Mol Psychiatry 13:833–857.

Phillips ML, Chase HW, Sheline YI, Etkin A, Almeida JR, Deckersbach T, Trivedi MH (2015) Identifying predictors, moderators, and mediators of antidepressant response in major depressive disorder: neuroimaging approaches. Am J Psychiatry 172:124–138.

Pilmeyer J, Huijbers W, Lamerichs R, Jansen JFA, Breeuwer M, Zinger S (2022) Functional MRI in major depressive disorder: A review of findings, limitations, and future prospects. J Neuroimaging 32:582–595.

Pimontel MA, Rindskopf D, Rutherford BR, Brown PJ, Roose SP, Sneed JR (2016) A Meta-Analysis of Executive Dysfunction and Antidepressant Treatment Response in Late-Life Depression. Am J Geriatr Psychiatry 24:31–41.

Poldrack RA (2000) Imaging brain plasticity: conceptual and methodological issues--a theoretical review. Neuroimage 12:1–13.

Rubinov M, Sporns O (2010) Complex network measures of brain connectivity: uses and interpretations. Neuroimage 52:1059–1069.

Ruhe HG, Booij J, Veltman DJ, Michel MC, Schene AH (2012) Successful pharmacologic treatment of major depressive disorder attenuates amygdala activation to negative facial expressions: A functional magnetic resonance imaging study. J Clin Psychiatry 73:451–459.

Santos M, Gold G, Kovari E, Herrmann FR, Hof PR, Bouras C, Giannakopoulos P (2010) Neuropathological analysis of lacunes and microvascular lesions in late-onset depression. Neuropathol Appl Neurobiol 36:661–672.

Sheline YI, Barch DM, Donnelly JM, Ollinger JM, Snyder AZ, Mintun MA (2001) Increased amygdala response to masked emotional faces in depressed subjects resolves with antidepressant treatment: An fmri study. Biol Psychiatry 50:651–658.

Shen X, Tokoglu F, Papademetris X, Constable RT (2013) Groupwise whole-brain parcellation from resting-state fMRI data for network node identification. Neuroimage 82:403–415.

Shi J, Malik J (2000) Normalized cuts and image segmentation. IEEE Transactions on pattern analysis and machine intelligence 22:888–905.

Shipp S (2004) The brain circuitry of attention. Trends Cogn Sci 8:223–230.

Siegle GJ, Thompson W, Carter CS, Steinhauer SR, Thase ME (2007) Increased amygdala and decreased dorsolateral prefrontal bold responses in unipolar depression: Related and independent features. Biol Psychiatry 61:198–209.

Skudlarski P, Jagannathan K, Calhoun VD, Hampson M, Skudlarska BA, Pearlson G (2008) Measuring brain connectivity: diffusion tensor imaging validates resting state temporal correlations. Neuroimage 43:554–561.

Smith GS, Workman CI, Kramer E, Hermann CR, Ginsberg R, Ma Y, Dhawan V, Chaly T, Eidelberg D (2011a) The relationship between the acute cerebral metabolic response to citalopram and chronic citalopram treatment outcome. Am J Geriatr Psychiatry 19:53–63.

Smith SM, Miller KL, Salimi-Khorshidi G, Webster M, Beckmann CF, Nichols TE, Ramsey JD, Woolrich MW (2011b) Network modelling methods for FMRI. Neuroimage 54:875–891.

Smith SM, Fox PT, Miller KL, Glahn DC, Fox PM, Mackay CE, Filippini N, Watkins KE, Toro R, Laird AR, Beckmann CF (2009) Correspondence of the brain’s functional architecture during activation and rest. Proc Natl Acad Sci U S A 106:13040–13045.

Speer AM, Kimbrell TA, Wassermann EM, J DR, Willis MW, Herscovitch P, Post RM (2000) Opposite effects of high and low frequency rtms on regional brain activity in depressed patients. Biol Psychiatry 48:1133–1141.

Stanley ML, Moussa MN, Paolini BM, Lyday RG, Burdette JH, Laurienti PJ (2013) Defining nodes in complex brain networks. Front Comput Neurosci 7:169.

Taylor SF, Liberzon I (2007) Neural correlates of emotion regulation in psychopathology. Trends Cogn Sci 11:413–418.

Tong Y, Hocke LM, Frederick BB (2019) Low Frequency Systemic Hemodynamic “Noise” in Resting State BOLD fMRI: Characteristics, Causes, Implications, Mitigation Strategies, and Applications. Front Neurosci 13:787.

Vos T, Barber RM, Bell B, Bertozzi-Villa A, Biryukov S, Bolliger I, Charlson F, Davis A, Degenhardt L, Dicker D (2015) Global, regional, and national incidence, prevalence, and years lived with disability for 301 acute and chronic diseases and injuries in 188 countries, 1990–2013: a systematic analysis for the Global Burden of Disease Study 2013. The lancet 386:743–800.

Wig GS, Schlaggar BL, Petersen SE (2011) Concepts and principles in the analysis of brain networks. Ann N Y Acad Sci 1224:126–146.

Wu J, Buchsbaum MS, Gillin JC, Tang C, Cadwell S, Wiegand M, Najafi A, Klein E, Hazen K, Bunney WE, Jr., Fallon JH, Keator D (1999) Prediction of antidepressant effects of sleep deprivation by metabolic rates in the ventral anterior cingulate and medial prefrontal cortex. Am J Psychiatry 156:1149–1158.

Xue SW, Li D, Weng XC, Northoff G, Li DW (2014) Different neural manifestations of two slow frequency bands in resting functional magnetic resonance imaging: A systemic survey at regional, interregional, and network levels. Brain Connect 4:242–255.

